# Peritraumatic C-reactive protein levels predict pain outcomes following traumatic stress exposure in a sex-dependent manner

**DOI:** 10.1101/2024.12.03.24318221

**Authors:** Lauren A. McKibben, Miranda N. Layne, Liz Marie Albertorio-Sáez, Ying Zhao, Erica M. Branham, Stacey L. House, Francesca L. Beaudoin, Xinming An, Jennifer S. Stevens, Thomas C. Neylan, Gari D. Clifford, Laura T. Germine, Kenneth A. Bollen, Scott L. Rauch, John P. Haran, Alan B. Storrow, Christopher Lewandowski, Paul I. Musey, Phyllis L. Hendry, Sophia Sheikh, Christopher W. Jones, Brittany E. Punches, Robert A. Swor, Lauren A. Hudak, Jose L. Pascual, Mark J. Seamon, Elizabeth M. Datner, David A. Peak, Roland C. Merchant, Robert M. Domeier, Niels K. Rathlev, Brian J. O’Neil, Leon D. Sanchez, Steven E. Bruce, John F. Sheridan, Steven E. Harte, Ronald C. Kessler, Karestan C. Koenen, Kerry J. Ressler, Samuel A. McLean, Sarah D. Linnstaedt

**Author notes:** <u>Manuscript correspondence</u>: Sarah D. Linnstaedt, PhD, University of North Carolina at Chapel Hill Campus Box #7010, Chapel Hill, NC 27599-7010.

## Abstract

**Background:** Chronic pain following traumatic stress exposure (TSE) is common. Increasing evidence suggests inflammatory/immune mechanisms are induced by TSE, play a key role in the recovery process versus development of post-TSE chronic pain, and are sex specific. In this study, we tested the hypothesis that the inflammatory marker C-reactive protein (CRP) is associated with chronic pain after TSE in a sex-specific manner.

**Methods:** We utilized blood-plasma samples and pain questionnaire data from men (n=99) and (n=223) women enrolled in *AURORA*, a multi-site emergency department (ED)-based longitudinal study of TSE survivors. We measured CRP using Ella/ELISA from plasma samples collected in the ED (‘peritraumatic CRP’, n=322) and six months following TSE (n=322). Repeated measures mixed-effects models were used to assess the relationship between peritraumatic CRP and post-TSE chronic pain.

**Results:** Peritraumatic CRP levels significantly predicted post-TSE chronic pain, such that higher levels of CRP were associated with lower levels of pain over time following TSE, but only in men (men:β=-0.24, *p*=0.037; women:β=0.05, *p*=0.470). By six months, circulating CRP levels had decreased by more than half in men, but maintained similar levels in women (t(290)=1.926, *p*=0.055). More men with a decrease in CRP levels had decreasing pain over time versus women (men:83% women:65%; Z=2.21, *p*=0.027).

**Conclusions:** In men but not women, we found circulating peritraumatic CRP levels predict chronic pain outcomes following TSE and resolution of CRP levels in men over time might be associated with increased pain recovery. Further studies are needed to validate these results.

**Summary:** Peritraumatic circulating CRP levels predicted pain recovery in men following TSE and decreases in CRP levels over time were greater in men compared to women.

## Introduction

Chronic posttraumatic musculoskeletal pain is a common long-term consequence of traumatic stress exposure (TSE).[54] For example, among individuals presenting to the emergency department for care following motor vehicle collision–a common life-threatening TSE in civilians–the prevalence of persistent moderate or severe pain exceeds 60%.[6] Chronic pain following TSE results in substantial suffering, functional interference, greater individual and societal costs, and a heightened risk of substance abuse and co-occurring neuropsychiatric symptoms.[5; 7–9; 23; 33; 36; 42; 47; 50; 51; 59] Consistent with other pain types,[20; 43; 44; 52] women have a disproportionate burden of post-TSE pain.[6]

There is a pressing need for research focused on improving understanding of molecular predictors and mechanisms of post-TSE pain development. Such research could lead to improved risk prediction tools and/or provide insights to inform the development of new preventive or treatment interventions. Increasing evidence suggests immune-related mechanisms play an important role in the pathogenesis of chronic pain after TSE and inflammatory mediators are a valuable area for study.[26; 41; 53]

One such inflammatory mediator is C-reactive protein (CRP). In healthy individuals, CRP levels in the blood are low but can rise dramatically in response to environmental stimuli such as TSE, injury, surgery, and infection.[12] Increases in CRP levels are often triggered by inflammatory cytokines such as IL-6, IL-1β, and TNF-α. [12; 46] CRP subsequently activates additional components of the immune system, particularly the innate immune system, facilitating the recruitment of immune cells to sites of inflammation.[37] Elevated CRP levels have been associated cross-sectionally with persistent pain states[1; 17; 18; 28] and longitudinally with chronic pain and associated recovery processes following acute injury.[25; 30] However, to date, no studies have examined the relationship between peritraumatic and longitudinal CRP levels and chronic pain development following TSE.

Available evidence suggests immune processes, chronic pain states, and the interplay between them often differs between the sexes.[4; 20; 32; 34; 35; 43; 48; 55] For instance, in male mice pain signaling in the spinal cord is mediated by microglia, whereas in female mice T cells are responsible for this signaling.[58] Research has also shown that TLR4, an activator of pro-inflammatory cytokines such as IL-6, IL-1β, and TNF-α, mediates hypersensitivity in male but not female mice.[57] Furthermore, CRP levels have been found to be sex-dependently associated with disease states related to chronic pain following TSE.[13; 15; 29] Given these significant sex differences in immune and pain mechanisms, independent analyses according to sex are essential for identifying key variations in the relationship between CRP and chronic pain.

In this study we used blood samples and data from motor vehicle collision survivors in the AURORA study, an emergency department (ED)-based, longitudinal, observational study of TSE survivors, to measure plasma CRP levels in men (n=99) and women (n=223) both in the immediate aftermath of TSE and six months later. We assessed the relationship between CRP levels and pain phenotypes. Our primary hypothesis was that peritraumatic circulating CRP levels would predict chronic pain outcomes in a sex-dependent manner. In secondary analyses, we assessed cross-sectional relationships between CRP and pain and whether changes in CRP levels over time were associated with pain recovery phenotypes in both men and women.

## Methods

### Study design and participants

The Advancing Understanding of RecOvery afteR traumA (AURORA) study was a longitudinal observational cohort study designed to improve the prevention, diagnosis, and treatment of adverse posttraumatic neuropsychiatric sequelae following trauma. Full eligibility criteria and study procedures have been previously described,[40] and are summarized as follows: participants aged 18-75 years who presented within 72 hours of TSE to one of 23 participating ED sites were screened for study eligibility, enrolled, and followed serially for one year.[40] Data collected over the course of follow-up and relevant to the current manuscript includes self-report questionnaires and biological samples. The study was approved by the Institutional Review Boards of all participating institutions, and all study participants provided written informed consent.

A subset from the overall AURORA cohort (n=2,943) was selected for this analysis based on availability of CRP data. To reduce heterogeneity we restricted our sample to individuals who had been involved in a motor vehicle collision as the reason for their ED visit, the predominant trauma type in the AURORA cohort (n=2,194, 74.5%). In total, n=322 participants with both ED (i.e ‘peritraumatic’) and 6-month blood samples were included in the current analyses. This subset was similar to the full cohort with respect to age, sex, race, and distribution of adverse outcomes. All participants assigned male and female sex at birth identified as men and women at the time of enrollment, so a binary categorization representing both sex and gender was used throughout all analyses.

### Pain assessments and outcome definition

Participants were asked to rate their current pain severity in the ED, over the past two weeks (at the 2-week time point), and in the past thirty days (at the eight-week, three-month, and six-month time points) via the validated 0-10 numeric rating scale (NRS: 0 (no pain) to 10 (maximum possible pain)).[16; 19; 31]

### Blood plasma collection

Blood samples were collected into 10-mL Vacutainer EDTA tubes (Beckton Dickinson, Franklin Lakes, NJ, USA, Catalog #366643) at baseline in the ED and six-months post-TSE. Within 30 minutes of collection, EDTA tubes were centrifuged (1,500g for 15 minutes), the plasma layer was removed and aliquoted (250μl per aliquot), and the individual aliquots were frozen at −20°C to −80°C at participating study sites. All samples were shipped in batches to the University of North Carolina at Chapel Hill BioSpecimen Processing Facility on dry ice and stored at −80°C until use.

### Quantification of CRP from human plasma samples

Human CRP levels were measured using a Simple Plex assay from the solid-phase sandwich antibody-based Ella System (Protein Simple, Biotechne, Minneapolis, MN, USA). This system has mostly automated the Enzyme-linked Immunosorbent Assay (ELISA) protocol and can measure levels of CRP between 32.8 - 50,000 pg/mL. In brief, 10μL of blood plasma from each sample were thawed on the day of analysis. Samples were diluted between 1:500 and 1:2000 according to manufacturer recommendations and based on serial dilution tests. Concentrations for each participant sample were calculated using the Simple Plex Runner 3.7.2.0 based on the assay’s standard curve. All data were converted to mg/dL to better enable comparisons with prior research. The intra- and inter-assay coefficient of variation between samples and assay plates was ≤ 10%. A few samples with higher variation were repeated or excluded from the analysis. CRP values in this cohort ranged from 0.001 to 8.285 mg/dL.

### Statistical analyses

Sociodemographic characteristics were summarized using standard descriptive statistics. CRP levels were determined to be non-normally distributed via the Shapiro-Wilk test[56]; therefore, a Box-Cox analysis[49] was performed and based on the λ value and a log transformation was identified as the optimal transformation. Therefore, CRP levels were log_2_ transformed. Participants with missing data were excluded from individual statistical tests on a pairwise (i.e. analysis-by-analysis) basis. CRP levels at the ED and six months following TSE were compared across men and women using t-tests. Repeated measures mixed models were used to assess the relationship between CRP levels and longitudinal pain outcomes separately in women and men, controlling for participant age, self-reported race/ethnicity, education level, ED pain levels, and CRP technical batches. Because BMI data was only available for a substantially smaller subset of the participants, we also ran our primary analysis (i.e. ED CRP and longitudinal pain severity) using only this subset of participants and including BMI as a covariate. Predicted pain values for each participant were derived from fully adjusted models and used for data visualization purposes. In data visualization graphs divided into ‘high CRP’ and ‘low CRP’ groups, participants were separated using the median level of CRP as a cutoff.

## Results

### Study design

Data and samples collected from AURORA study[40] participants who experienced a motor vehicle collision TSE were used in the present analysis (**Figure 1**). For this analysis, participants reported to the ED within 72 hours of their TSE, provided a blood sample, and reported their current pain intensity. They also reported pain intensity at three intervals following their TSE (eight weeks, three months, and six months). At six months following TSE, each participant provided a second blood sample. Using both ED and six-month blood samples, CRP was assayed using a semi-automated ELISA technology and was analyzed in relation to pain intensity.

**Figure 1.**
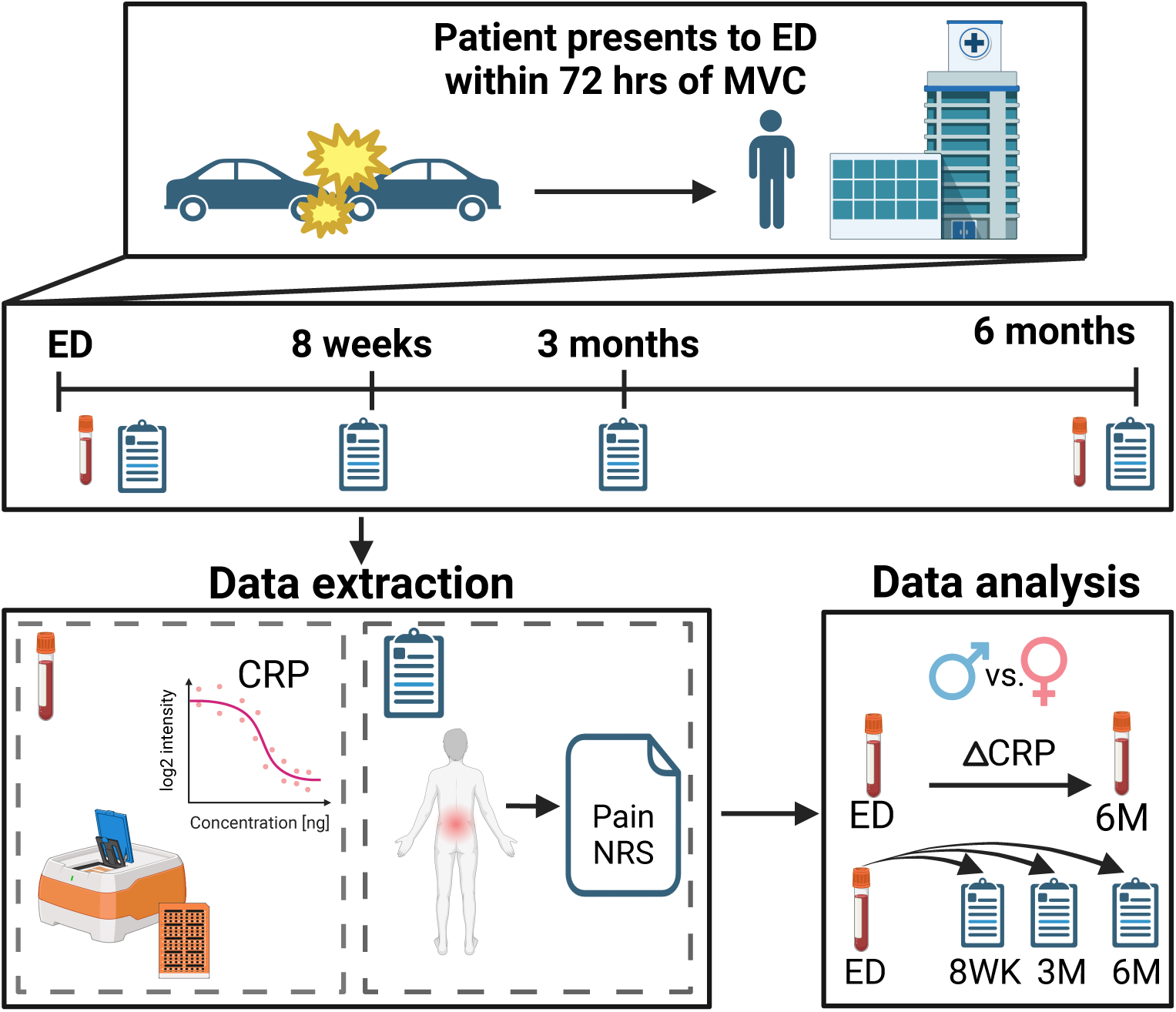
Overview of data collection, extraction, and analysis. This research study utilizes data from the AURORA cohort, a human cohort study comprised of participants who have experienced a motor vehicle collision traumatic stress exposure (TSE) to test the relationship between circulating C-reactive protein (CRP) levels and post-TSE chronic pain. The AURORA study enrolled individuals who reported to the emergency department (ED) within 72 hours of TSE and provided a blood sample. Relevant to the current study, each patient reported their overall pain intensity in the ED and at three intervals following their TSE (eight weeks, three months, and six months). A second blood-plasma sample was collected at six months. CRP was assayed in both the ED and six-month plasma samples using Ella/ELISA technology. The data was then analyzed by measuring the relationship between CRP and pain at each timepoint individually, the relationship between circulating peritraumatic CRP levels and longitudinal pain intensity, and the change in CRP from ED to month six, all in men and women separately to allow for the identification of sex-specific effects. Created in BioRender. Layne, M. (2025) https://BioRender.com/f20t989

### AURORA study participant characteristics

This study included 322 participants who were selected based on having experienced a motor vehicle collision trauma and had available blood-plasma. These participants were similar to the full AURORA cohort as based on key characteristics (e.g. age, sex, outcome distributions). Baseline characteristics of these participants are shown in **Table 1**. Most individuals were under 40 years of age, were either non-Hispanic Black (60.6%) or non-Hispanic White (39.4%), were women (69.3%), and had at least some post-secondary education (67.7%). In the following analyses, women and men were studied separately due to the known sex differences in CRP association with various disease states[10; 13; 15; 29] and known sex differences in chronic posttraumatic musculoskeletal pain vulnerability[6; 24; 45]

**Table 1.** Baseline characteristics of AURORA study participants included in current analyses.

|  | **Men**<br>**(n=99)** | **Women**<br>**(n=223)** | **Combined**<br>**Cohort (n=322)** |
| --- | --- | --- | --- |
| Age, years, mean (SD) | 38.6 (14.8) | 40.5 (13.6) | 39.9 (14.0) |
| BMI, mean (SD) | 30.2 (9.8) | 32.5 (8.6) | 31.9 (9.0) |
| Education, n (%) |  |  |  |
| Less than high school graduate | 14 (14.1%) | 21 (9.4%) | 35 (10.9%) |
| High school graduate | 25 (25.3%) | 44 (19.7%) | 69 (21.4%) |
| Some college | 43 (43.4%) | 98 (43.9%) | 141(43.8%) |
| College graduate | 17 (17.2%) | 60 (26.9%) | 77 (23.9%) |
| Ethnicity, n (%) |  |  |  |
| Non-Hispanic White | 43 (43.4) | 84 (37.7) | 127 (39.4) |
| Non-Hispanic Black | 56 (56.6) | 139 (62.3) | 195 (60.6) |
| ADI, mean (SD) | 62.4 (32.4) | 65.4 (28.2) | 64.5 (29.5) |
| Pain severity<sup>a</sup> in the ED, mean (SD) | 6.26 (2.87) | 6.34 (2.52) | 6.32 (2.63) |
| Previous traumatic events<sup>b</sup>, mean (SD) | 3.29 (2.68) | 3.33 (2.49) | 3.32 (2.54) |

<sup>a</sup> - Pain severity measured via the 0-10 numeric rating scale
<sup>b</sup> - Previous traumatic events assessed via the life events checklist
SD = standard deviation, BMI = body mass index, ED = emergency department, ADI = Area Deprivation Index

### Women recover more slowly than men following motor vehicle collision trauma

We previously showed that women in the AURORA study are more likely to develop chronic pain following TSE than men.[6] We wanted to confirm this association because we were using a subset of the AURORA dataset. We first used repeated measures mixed models adjusted for participant age, education, pain intensity reported in the ED, and race/ethnicity. As shown in **Figure 2**, women and men had similar acute pain levels, but women recovered more slowly over time than men (**Figure 2**, β=0.579, *p*=0.048). When assessing if there were sex differences in pain levels at each follow-up timepoint, we found that women had statistically significantly more pain six months following TSE compared to men (β=-1.327, *p*<0.001), but pain level difference by sex did not reach statistical significance at the eight-week or three-month post-TSE timepoints (*p*>0.05).

**Figure 2.**
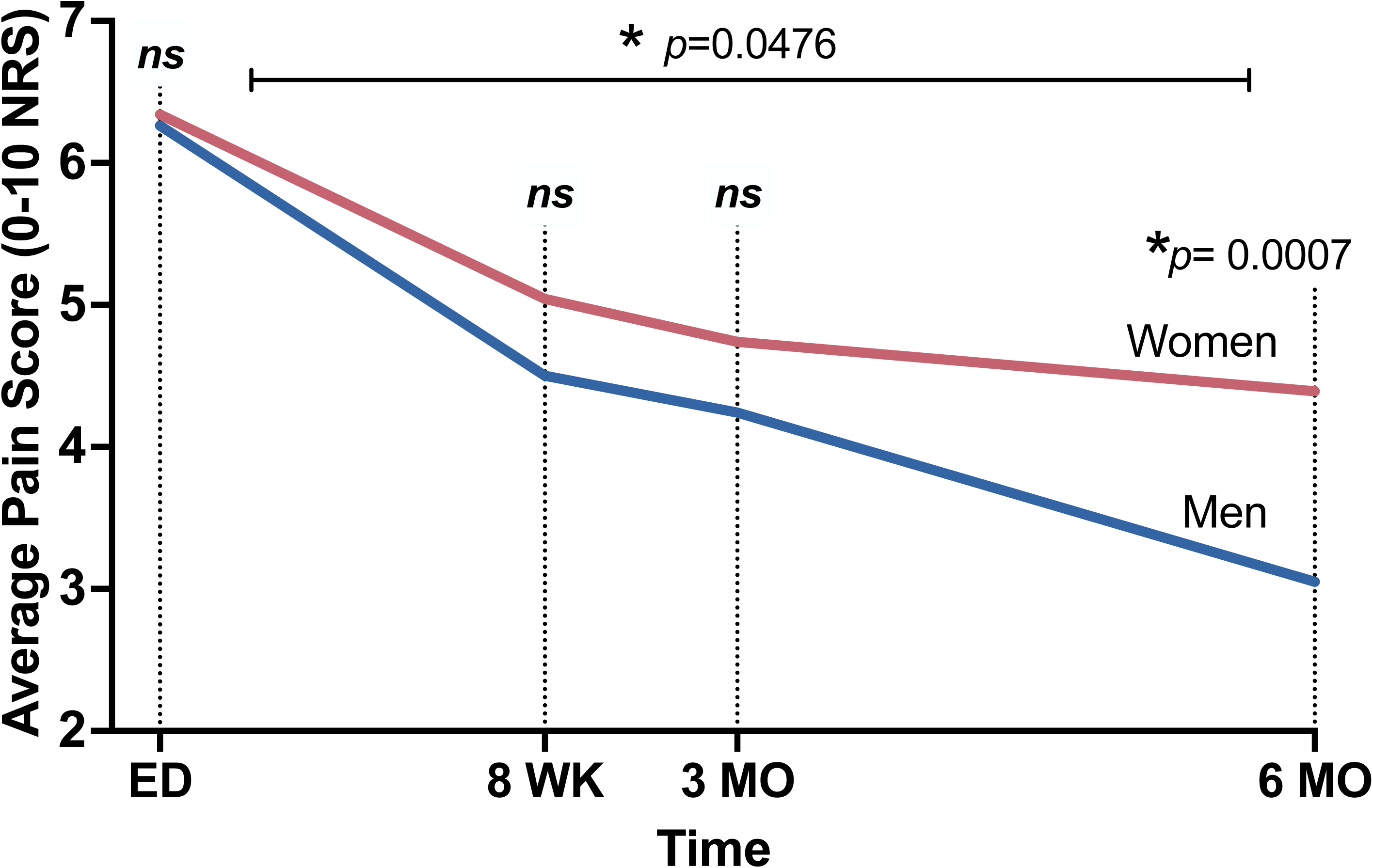
Chronic posttraumatic pain trajectories following traumatic stress exposure in men (n=99) and women (n=220) from the AURORA study that had corresponding blood plasma samples and were used for the current analyses. Pain intensity at three timepoints following enrollment were graphed relative to ED pain levels. Red line: women and blue line: men. *p<0.05 as indicated. Average pain scores were assessed via verbal numeric rating scales (NRS). ED: emergency department, WK: week, MO: month, ns: non-significant.

### Circulating CRP levels in the early aftermath of TSE do not differ between women and men and are not associated with acute pain levels in either sex

Before evaluating the relationship between circulating CRP levels and longitudinal pain outcomes following TSE, we first assessed the relationship between time-matched CRP and pain scores in the acute aftermath of TSE. Mean circulating CRP levels in the ED were consistent with moderately elevated levels of CRP in adults (1.26±1.7mg/dL).[2; 46] Using a t-test, CRP levels at the time of TSE did not differ between men and women (men:1.26±2.0mg/dL vs women: 1.26±1.6mg/dL t(320)=0.553, *p*=0.581; **Figure 3a**). However, of note, 38.6% of women had CRP levels above the normal threshold (i.e. 1.0mg/dL) and 29.3% of men had CRP levels above the normal threshold. In linear regression models adjusted for previously mentioned demographic characteristics and batch effects, we found peritraumatic CRP levels measured in the ED were not associated with pain severity reported in the ED in men or women (men: β=0.052, *p*=0.676; women: β=0.102, *p*=0.120; **Figure 3b**).

**Figure 3.**
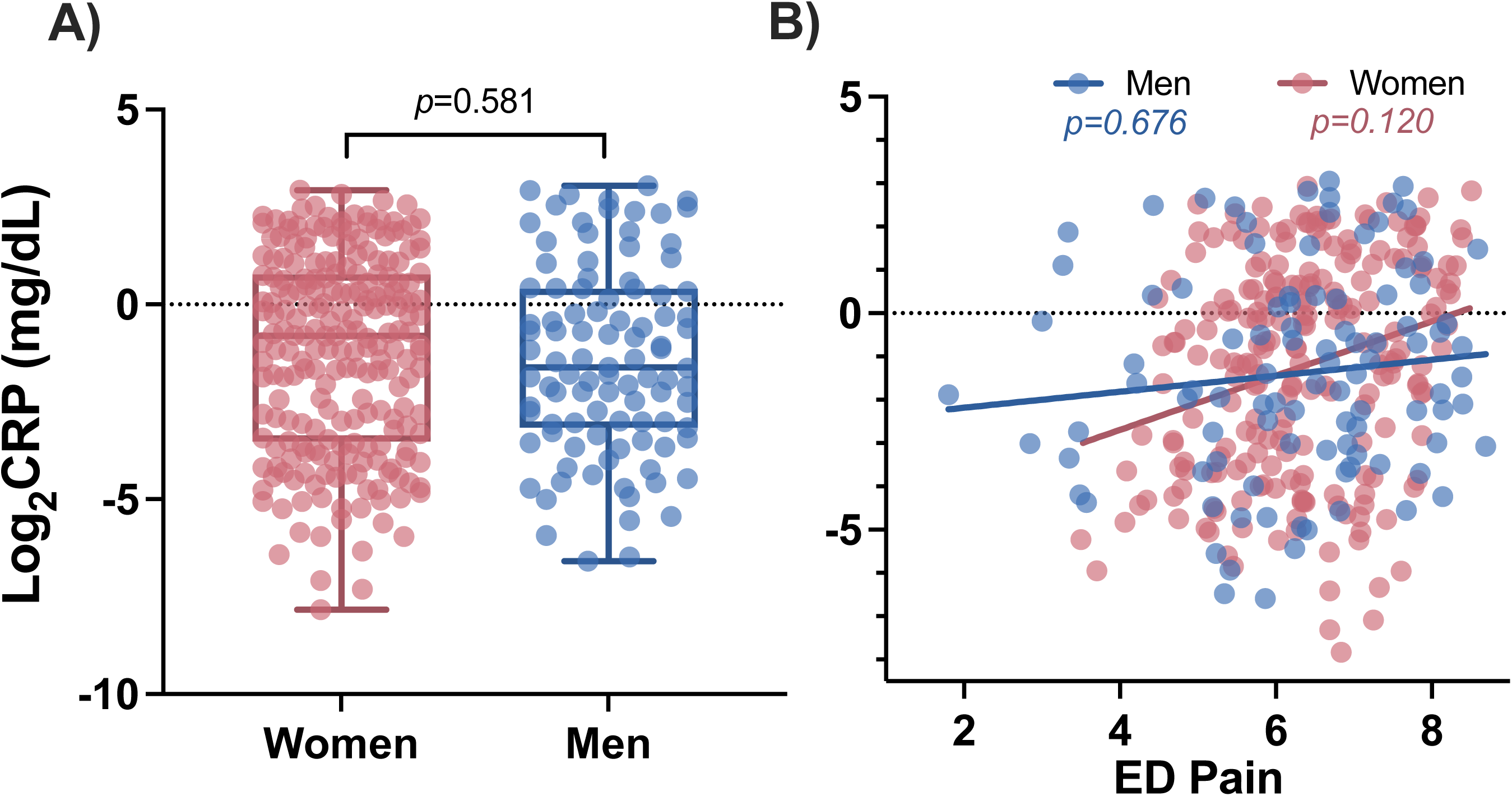
Peritraumatic circulating CRP levels are similar between women and men in the immediate aftermath of traumatic stress exposure (i.e. at the ED timepoint) and are not associated with levels of acute pain assessed in the ED. A) Circulating Log_2_CRP levels in the ED and B) the relationship between circulating CRP levels from the ED timepoint and pain levels that were also assessed at the ED timepoint, are shown. CRP: C-reactive protein, ED: emergency department, mg/dL: milligrams per deciliter.

### Peritraumatic circulating CRP levels predict post-TSE chronic pain outcomes in men but not in women

Our primary hypothesis was that peritraumatic circulating CRP levels would predict pain levels over time following TSE. As shown in (**Table 2**), we found that peritraumatic CRP levels predicted longitudinal pain outcomes in men but not in women (men: β=-0.24, *p*=0.037; women:β=0.05, *p*=0.470). Statistics corresponding to all covariates are shown in ***Supplementary Table 1***. Due to previously identified relationships between BMI and CRP,[14] we also tested this relationship with BMI included as a covariate. The addition of this covariate had little influence on the effect size of the relationship between CRP and longitudinal pain outcomes in men, but it reduced statistical significance (men: β=-0.23, *p*=0.110; women:β=0.04, *p*=0.643, ***Supplementary Table 2***). Because our sample size was substantially reduced in this analysis due to missing BMI data (n=197 women and n=78 men), we also tested our original model without the BMI covariate in this reduced cohort (***Supplementary Table 3***). Directly comparing models in this reduced cohort, we found little difference between including BMI as a predictor or not (with BMI: women: β=0.04, *p*=0.643; men: β=-0.23, *p*=0.110; without BMI: women:β=0.07, *p*=0.346; men: β=-0.23, *p*=0.096), and BMI itself was not a predictor of chronic pain in either men or women (men: β=-0.001, *p*=0.987; women:β=0.03, *p*=0.392).

**Table 2.**
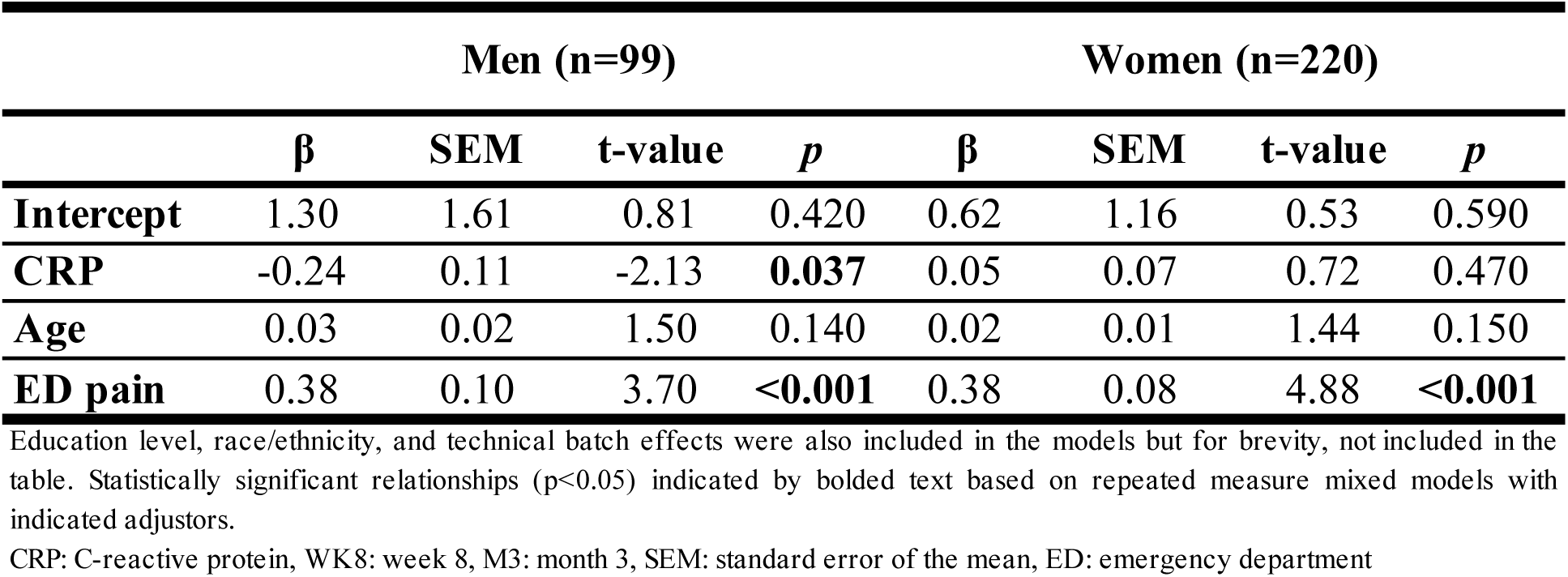
Relationship between peritraumatic circulating CRP levels and posttraumatic musculoskeletal pain (WK8, M3, M6) following traumatic stress exposure in men and women (n=319)

In our analyses examining the relationship between ED CRP levels and pain at each follow-up timepoint in men and women, controlling for the same demographic and assay variables as above, we found ED CRP levels statistically significantly predicted lower pain severity three months after TSE, but only in men (3-month:β=-0.32, *p*=0.0186) and not at eight weeks or six months in men (8-week:β=-0.23, *p*=0.096; 6-month:β=-0.21, *p*=0.181). In women, ED CRP levels were not associated with pain severity at any timepoint (*p*>0.05). To visualize the association between peritraumatic CRP levels and longitudinal pain outcomes, we divided participants into subgroups based on sex and peritraumatic CRP level (levels above vs. below the median). **Figure 4** shows the average pain severity at each post-TSE timepoint within these subgroups.

**Figure 4.**
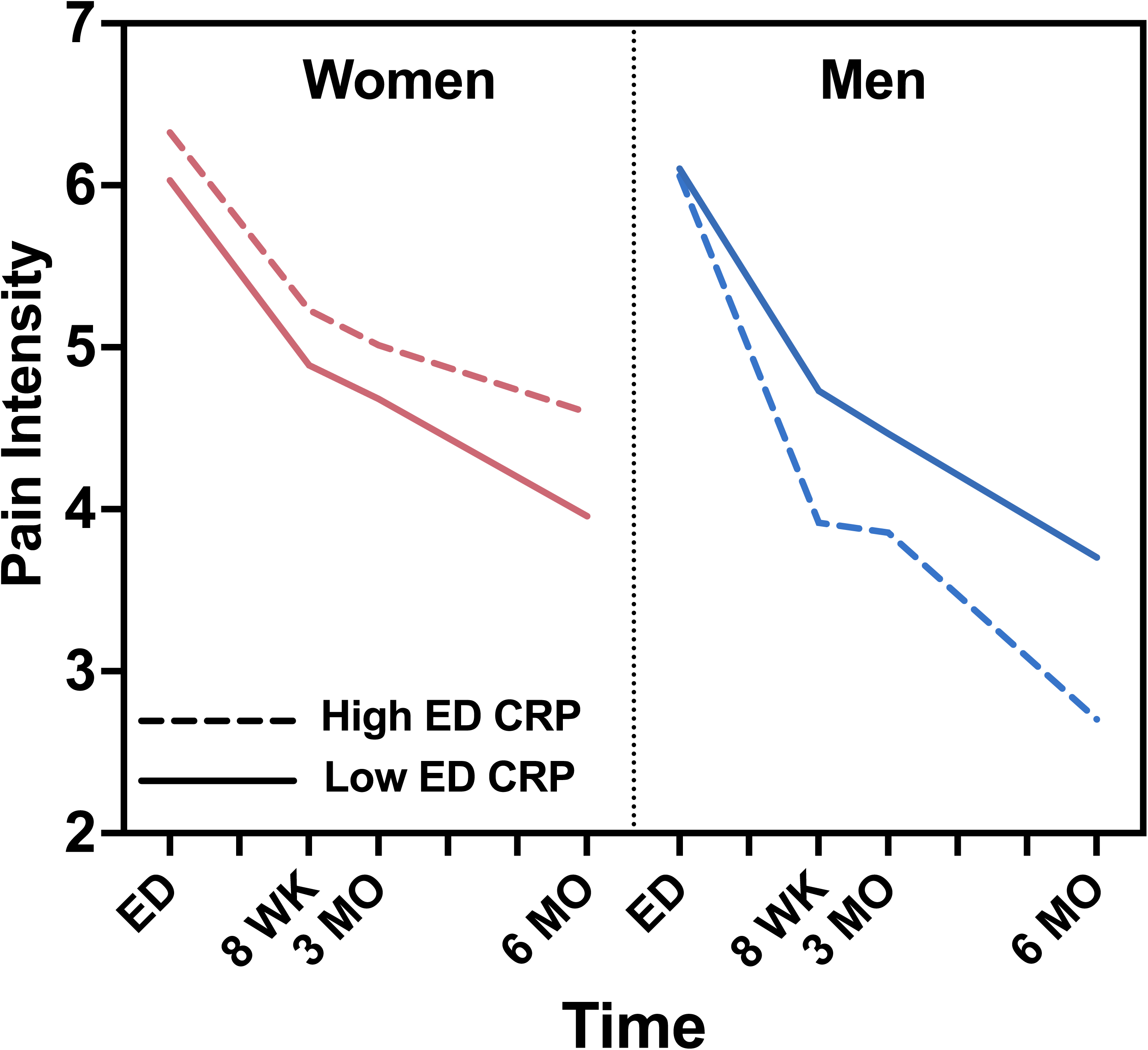
Average predicted pain intensity values in women and men with high and low peritraumatic CRP levels. Peritraumatic circulating CRP levels predicted pain outcomes in men but not women such that men with high CRP at the time of traumatic stress exposure have lower levels of pain longitudinally following TSE (men β=-0.24, *p*=0.037; women:β=0.05, *p*=0.470). Predicted pain values at each timepoint were extracted, corrected for all covariates except BMI, and the average value is shown for each group. Analyses were conducted using continuous CRP. Participants were split into high and low levels based on median CRP for visualization purposes. ED: emergency department, CRP: C-reactive protein, WK: week, MO: month, TSE: traumatic stress exposure. Pain intensity was based on the verbal numeric rating scale (0-10 NRS)

### CRP levels, sex, and pain 6 months after TSE

We next explored the relationship between CRP, sex, and pain six months after TSE. At six months, men had almost 2x less circulating CRP compared to women, though the difference was only statistically significant at the trend level (men: 0.53±0.8mg/dL vs women: 1.02±1.4mg/dL; t(290)=1.926, *p*=0.055; **Figure 5a**). At this six-month timepoint, only 15.2% of men had CRP levels above the normal threshold while 29.6% of women had CRP levels above the normal threshold (i.e. above 1.0mg/dL). We next assessed whether CRP levels were associated with pain levels at the six-month timepoint and found a stronger relationship in men than in women, but the relationship was not statistically significant in either sex (men: β=-0.29, *p*=0.122; women: β=-0.01, *p*=0.943; **Figure 5b**). Additionally controlling for ED pain in this model did not change the statistical outcome (men: β=-0.28, *p*=0.146; women: β=-0.04, *p*=0.694).

**Figure 5.**
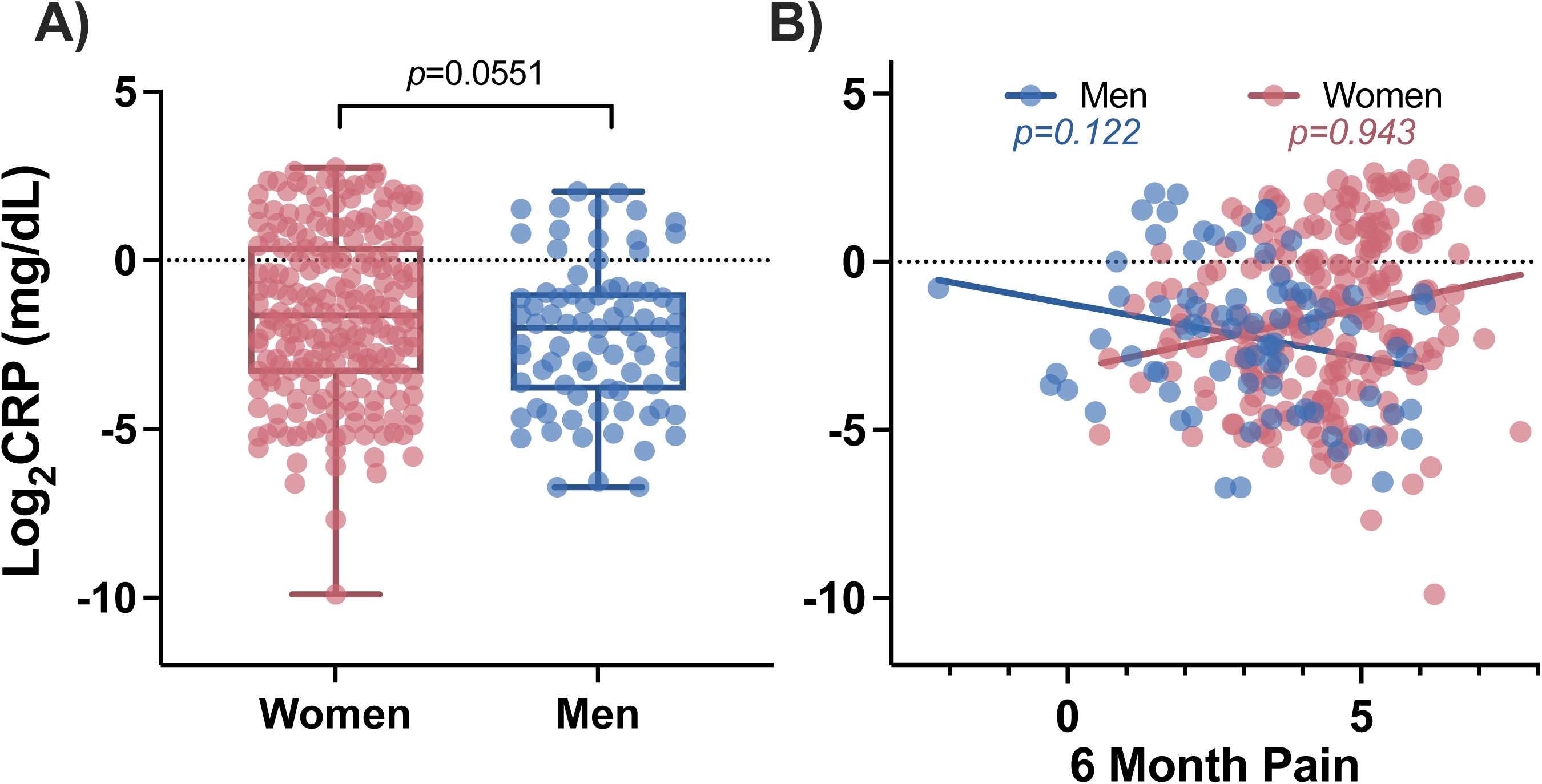
Relationship between circulating CRP levels measured in blood plasma samples collected six months following TSE and six-month pain levels in women and men. A) Log_2_CRP levels in men and women six months following TSE and B) the relationship between CRP levels at the six-month timepoint and pain levels that were also assessed six months following TSE, are shown. CRP: C-reactive protein, TSE: traumatic stress exposure, mg/dL: milligrams per deciliter.

### Changes in CRP levels over time in men vs. women

Decreasing CRP levels over time were more common in men than increasing levels (61% decreasing, 39% increasing; *Z* = 2.98, *p* = 0.003, **Figure 6a**). In contrast, in women, there was no difference in the percentage of women with decreasing versus increasing levels of CRP (53% decreasing, 47% increasing; Z=1.23, *p*=0.218). Further, when comparing the magnitude of decreasing levels of CRP in men vs women, men exhibited a greater drop in CRP levels between the early post-TSE timepoint and the six-month timepoint (mean change in men=-1.532, mean change in women= −1.085; t(176)=1.986, *p*=0.049; **Figure 6b**).

**Figure 6.**
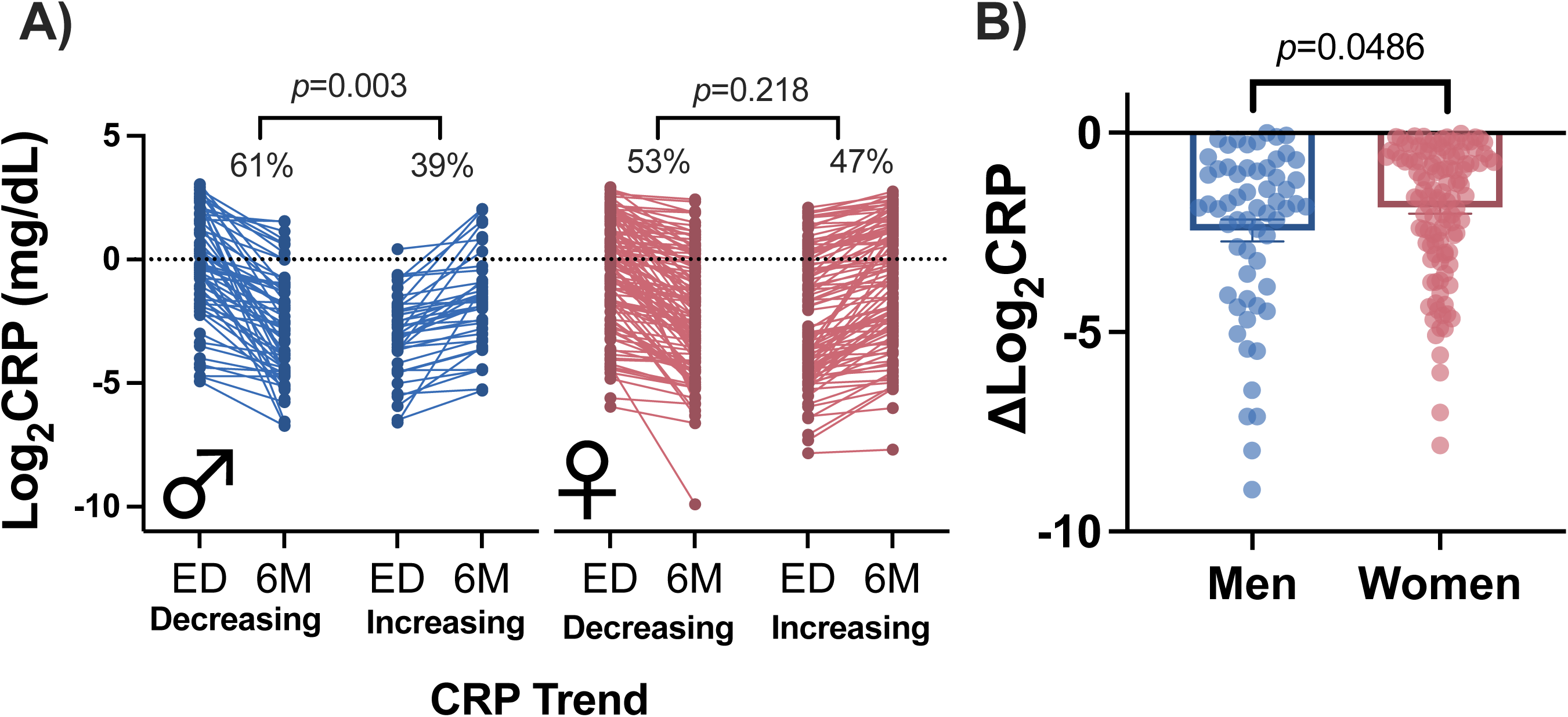
CRP levels from ED to six months following TSE for men and women. **A)** The percentage of women and men with decreasing and increasing CRP levels are shown. **B)** The average magnitude decrease in CRP is shown for men and women who have decreasing CRP levels. CRP: C-reactive protein, ED: emergency department, 6M: 6 months, TSE: traumatic stress exposure.

To examine whether the decrease in CRP levels between the ED and six-month timepoints in men were associated with decreased levels of pain in men, we performed chi-square analysis to determine whether there was overlap between men with decreasing pain levels and those with decreasing CRP (as well as increasing pain and increasing CRP)(***Supplementary Table 4***). However, we did not find a statistically significant relationship in men (χ^2^=4.32, *p*=0.115). Similarly, we did not find a statistically significant relationship in women (χ^2^=1.19, *p*=0.552). However, we found that more men with a decrease in CRP levels had decreasing pain over time versus women (men:83% women:65%; Z=2.21, *p*=0.027).

## Discussion

This study examined the relationship between plasma C-reactive protein (CRP) levels, sex, and pain outcomes after TSE. Increased peritraumatic plasma CRP levels predicted pain resolution in men but not women. Unlike women, most men exhibited reductions in CRP levels over six months following TSE. Compared to women, these reductions in men often coincided with reductions in pain.

The results from our additional analyses indicated CRP levels measured at each cross-sectional time point (emergency department (ED) visit and six months later) were not associated with pain levels at those same time points. Notably, average CRP levels were slightly elevated in both sexes during the ED visit based on standard pre-defined cutoffs[46] but dropped to normal levels in men after more than a two-fold decrease over the six month period post-TSE. Collectively, these findings suggest CRP may serve as a predictor of pain recovery in men.

Further studies are needed to determine if CRP specifically, or inflammation more generally, plays a mechanistic role in the resolution of pain following TSE. Recent evidence suggests that an acute inflammatory response is important for pain resolution. For instance, a recent study found a neutrophil-driven immune response was essential to the resolution of low back pain, and the administration of anti-inflammatory agents inhibited pain recovery.[53] Other research suggested the role of inflammation-reducing molecular mediators (such as resolvins and protectins) in facilitating pain resolution over time.[22; 27; 61] Further, in a recent animal study conducted by our group, we examined gene expression profiles following severe acute stress and found immune system activation within 24 to 72 hours post-stress.[39] This study, in which the animals fully recovered over the subsequent several days, suggests immune activation is a natural response to TSE and assists in the natural recovery process. These findings align with other studies indicating that stress-immune processes are natural mediators of the recovery process following TSE and suggests impairments to this process (e.g. persistent CRP levels over time in women) increase risk for pain persistence. Future research assessing the role CRP plays in recovery from TSE, including sex-specific effects on pain resolution, are warranted.

We showed here that, consistent with a larger cohort of TSE survivors [6], women had higher pain levels six months following TSE versus men. Further studies are needed to determine if the observed sex differences in CRP levels contribute to these differences in clinical outcomes. While the reasons for these sex differences are not known, it is possible sex hormones could account for differences in CRP levels in men vs women, as previous research has shown sex hormones can influence CRP levels[11; 60]. Future studies should explore this possibility.

The mechanisms through which CRP might influence pain outcomes after TSE are not known. However, previous research from other groups has proposed several possible mechanisms for CRP in post-TSE pathologies. First, persistently high CRP levels may be influencing pain perception by maintaining or exacerbating the inflammatory environment (via activation of complement or other immune cells), leading to tissue damage and/or continuous nociceptive input from the periphery to the central nervous system via direct sensitization of nociceptors by inflammatory mediators (e.g. PGE2, NGF).[3; 38; 46] In addition, other studies have suggested that CRP, via its ability to limit neutrophil chemotaxis and neutrophil production of reactive oxygen species, is involved not in the persistence of inflammation but rather in the resolution of inflammation.[21] Whether CRP is directly involved in the pathogenesis of post-TSE chronic pain or is a marker of inflammation and its persistence/resolution has yet to be determined.

Strengths of our study include the use of a longitudinal multi-ethnic cohort, sex-stratified analyses, CRP levels from paired longitudinal samples collected at the time of trauma exposure and six months following TSE, and adjustment for factors in our models that commonly confound such analyses. Several limitations should also be considered when interpreting our study results. First, and most importantly, these results have not been validated in a second cohort. Therefore, we do not know the generalizability of our findings. Second, related to the first limitation, the sample size is relatively small, limiting our ability to further stratify the population beyond sex-stratification, or to adjust for additional confounders. Third, due to the nature of the study design, we were not able to assess pre-trauma CRP levels to be able to identify whether TSEs cause an increase in circulating CRP. Fourth, CRP exists in both pentameric and monomeric forms, and differentiating between these forms can be an avenue for research aimed at understanding CRP’s full biological function, however this distinction was not explored in the current study.

In summary, we found that peritraumatic CRP predicts chronic pain development over the subsequent six months and CRP levels decreased in men but not women. Future studies should aim to validate these results in a larger cohort and identify causal mechanisms driving sex-dependent changes in CRP levels over time and their associations with chronic pain outcomes following TSE.

## Supporting information

Figure 1 Publication License

## Data Availability Statement

Data used in this manuscript is available for download from the UNC Dataverse at https://doi.org/10.15139/S3/EEPWIC. These data and all other data collected through the AURORA Project are available through the National Institute of Mental Health (NIMH) Data Archive (NDA) found here: https://nda.nih.gov/edit_collection.html?id=2526.

## Acknowledgements

The investigators wish to thank the trauma survivors participating in the AURORA Study. Their time and effort during a challenging period of their lives make our efforts to improve recovery for future trauma survivors possible. The authors would also like to acknowledge the University of North Carolina BioSpecimen Facility for the storage, accessioning and disbursement of biological samples. Research reported in this publication was supported by the National Institute of Arthritis and Musculoskeletal and Skin Diseases, the National Institute of Mental Health, the National Institute of Neurological Disorders and Stroke, and the National Institute of General Medical Science of the National Institutes of Health under Award Numbers U01MH110925 (McLean, Ressler, Kessler, and Koenen), K01AR071504 (Linnstaedt), R01AR081454 (Linnstaedt), R01NS118563 (Linnstaedt and McLean), and T32GM008450 (T32 fellow McKibben). Additional funding was provided by the Rita Allen Foundation (Linnstaedt), the US Army MRMC, One Mind, and The Mayday Fund. The content is solely the responsibility of the authors and does not necessarily represent the views of these funding agencies. This manuscript reflects the views of the authors and may not reflect the opinions or views of the NIH or of the Submitters submitting original data to NDA.

## References

[1] Afari N, Mostoufi S, Noonan C, Poeschla B, Succop A, Chopko L, Strachan E. C-reactive protein and pain sensitivity: findings from female twins. Ann Behav Med 2011;42(2):277–283.

[2] Ansar W, Ghosh S. C-reactive protein and the biology of disease. Immunol Res 2013;56(1):131–142.

[3] Barker PA, Mantyh P, Arendt-Nielsen L, Viktrup L, Tive L. Nerve Growth Factor Signaling and Its Contribution to Pain. J Pain Res 2020;13:1223–1241.

[4] Bartley EJ, Fillingim RB. Sex differences in pain: a brief review of clinical and experimental findings. Br J Anaesth 2013;111(1):52–58.

[5] Beaudoin FL, Gutman R, Merchant RC, Clark MA, Swor RA, Jones JS, Lee DC, Peak DA, Domeier RM, Rathlev NK. Persistent pain after motor vehicle collision: comparative effectiveness of opioids versus non-steroidal anti-inflammatory drugs prescribed from the emergency department--a propensity matched analysis. Pain 2017;158(2):289.

[6] Beaudoin FL, Kessler R, Hwang I, Lee S, Sampson N, An X, Ressler K, Koenen K, McLean S, Group AS. Pain after a motor vehicle crash: The role of socio demographics, crash characteristics and peri traumatic stress symptoms. European journal of pain 2021;25(5):1119–1136.

[7] Beaudoin FL, Straube S, Lopez J, Mello MJ, Baird J. Prescription opioid misuse among ED patients discharged with opioids. The American journal of emergency medicine 2014;32(6):580–585.

[8] Beck JG, Clapp JD. A different kind of comorbidity: Understanding posttraumatic stress disorder and chronic pain. Psychological Trauma: Theory, Research, Practice, and Policy 2011;3(2):101.

[9] Beckham JC, Crawford AL, Feldman ME, Kirby AC, Hertzberg MA, Davidson J, Moore SD. Chronic posttraumatic stress disorder and chronic pain in Vietnam combat veterans. Journal of psychosomatic research 1997;43(4):379–389.

[10] Berwal D, Branisteanu DD, Glickman M, Sagar A, Pilitsis JG. The sex-dependent impact of adipose tissue and inflammation on chronic pain - A cross-sectional study from the all of us research program. Cytokine 2024;179:156614.

[11] Bianchi VE. The anti-inflammatory effects of testosterone. Journal of the endocrine society 2019;3(1):91–107.

[12] Black S, Kushner I, Samols D. C-reactive Protein. J Biol Chem 2004;279(47):48487–48490.

[13] Chen B, Dong J, Guo W, Li T. Sex-specific associations between levels of high-sensitivity C-reactive protein and severity of depression: retrospective cross-sectional analysis of inpatients in China. BMC Psychiatry 2024;24(1):667.

[14] Choi J, Joseph L, Pilote L. Obesity and C reactive protein in various populations: a systematic review and meta analysis. Obesity reviews 2013;14(3):232–244.

[15] Dal Santo F, González-Blanco L, García-Álvarez L, de la Fuente-Tomás L, Velasco Á, Álvarez-Vázquez CM, Martínez-Cao C, Sáiz PA, García-Portilla MP, Bobes J. Cognitive impairment and C-reactive protein in clinically stable schizophrenia outpatients: a focus on sex differences. Sci Rep 2020;10(1):15963.

[16] Farrar JT, Young JP, Jr., LaMoreaux L, Werth JL, Poole MR. Clinical importance of changes in chronic pain intensity measured on an 11-point numerical pain rating scale. Pain 2001;94(2):149–158.

[17] Farrell SF, Armfield NR, Cabot PJ, Elphinston RA, Gray P, Minhas G, Collyer MR, Sterling M. C-Reactive Protein (CRP) is Associated With Chronic Pain Independently of Biopsychosocial Factors. J Pain 2024;25(2):476–496.

[18] Farrell SF, Sterling M, Klyne DM, Mustafa S, Campos AI, Kho PF, Lundberg M, Rentería ME, Ngo TT, Cuéllar-Partida G. Genetic impact of blood C-reactive protein levels on chronic spinal & widespread pain. Eur Spine J 2023;32(6):2078–2085.

[19] Ferreira-Valente MA, Pais-Ribeiro JL, Jensen MP. Validity of four pain intensity rating scales. Pain 2011;152(10):2399–2404.

[20] Fillingim RB, King CD, Ribeiro-Dasilva MC, Rahim-Williams B, Riley JL, 3rd. Sex, gender, and pain: a review of recent clinical and experimental findings. J Pain 2009;10(5):447–485.

[21] Friend SF, Nachnani R, Powell SB, Risbrough VB. C reactive protein: Marker of risk for post traumatic stress disorder and its potential for a mechanistic role in trauma response and recovery. European Journal of Neuroscience 2022;55(9-10):2297–2310.

[22] Fullerton JN, Gilroy DW. Resolution of inflammation: a new therapeutic frontier. Nature reviews Drug discovery 2016;15(8):551–567.

[23] Gatchel RJ, McGeary DD, McGeary CA, Lippe B. Interdisciplinary chronic pain management: past, present, and future. Am Psychol 2014;69(2):119–130.

[24] Hadlandsmyth K, Driscoll MA, Johnson NL, Mares JG, Mengeling MA, Thomas EBK, Norman SB, Lund BC. Veterans with chronic pain: Examining gender differences in pain type, overlap, and the impact of post-traumatic stress disorder. Eur J Pain 2024;28(8):1311–1319.

[25] Hashimoto K, Tsuji A, Takenaka S, Ohmura A, Ueki R, Noma H, Imamura M, Miyoshi Y, Kariya N, Tatara T, Hirose M. C-reactive Protein Level on Postoperative Day One is Associated with Chronic Postsurgical Pain After Mastectomy. Anesth Pain Med 2018;8(4):e79331.

[26] Ji R-R, Chamessian A, Zhang Y-Q. Pain regulation by non-neuronal cells and inflammation. Science 2016;354(6312):572–577.

[27] Ji R-R, Xu Z-Z, Gao Y-J. Emerging targets in neuroinflammation-driven chronic pain. Nature reviews Drug discovery 2014;13(7):533–548.

[28] Khamisy-Farah R, Fund E, Raibman-Spector S, Adawi M. Inflammatory Markers in the Diagnosis of Fibromyalgia. Isr Med Assoc J 2021;23(12):801–804.

[29] Khera A, McGuire DK, Murphy SA, Stanek HG, Das SR, Vongpatanasin W, Wians FH, Jr., Grundy SM, de Lemos JA. Race and gender differences in C-reactive protein levels. J Am Coll Cardiol 2005;46(3):464–469.

[30] Klyne DM, Barbe MF, Hodges PW. Relationship between systemic inflammation and recovery over 12 months after an acute episode of low back pain. Spine J 2022;22(2):214–225.

[31] Krebs EE, Carey TS, Weinberger M. Accuracy of the pain numeric rating scale as a screening test in primary care. J Gen Intern Med 2007;22(10):1453–1458.

[32] Levesque P, Desmeules C, Béchard L, Huot-Lavoie M, Demers MF, Roy MA, Deslauriers J. Sex-specific immune mechanisms in PTSD symptomatology and risk: A translational overview and perspectives. Brain Res Bull 2023;195:120–129.

[33] Lew HL, Otis JD, Tun C, Kerns RD, Clark ME, Cifu DX. Prevalence of chronic pain, posttraumatic stress disorder, and persistent postconcussive symptoms in OIF/OEF veterans: polytrauma clinical triad. Journal of Rehabilitation Research & Development 2009;46(6).

[34] Linnstaedt SD, Mauck MC, Son EY, Tungate AS, Pan Y, Rueckeis C, Yu S, Lechner M, Datner E, Cairns BA, Danza T, Velilla MA, Pearson C, Shupp JW, Smith DJ, McLean SA. Peritraumatic 17β-estradiol levels influence chronic posttraumatic pain outcomes. Pain 2021;162(12):2909–2916.

[35] Linnstaedt SD, Rueckeis CA, Riker KD, Pan Y, Wu A, Yu S, Wanstrath B, Gonzalez M, Harmon E, Green P, Chen CV, King T, Lewandowski C, Hendry PL, Pearson C, Kurz MC, Datner E, Velilla MA, Domeier R, Liberzon I, Mogil JS, Levine J, McLean SA. MicroRNA-19b predicts widespread pain and posttraumatic stress symptom risk in a sex-dependent manner following trauma exposure. Pain 2020;161(1):47–60.

[36] Lobo JJ, McLean SA, Tungate AS, Peak DA, Swor RA, Rathlev NK, Hendry PL, Linnstaedt SD. Polygenic risk scoring to assess genetic overlap and protective factors influencing posttraumatic stress, depression, and chronic pain after motor vehicle collision trauma. Translational Psychiatry 2021;11(1):359.

[37] Ma YJ, Garred P. Pentraxins in Complement Activation and Regulation. Front Immunol 2018;9:3046.

[38] Macphail K. C-reactive protein, chronic low back pain and, diet and lifestyle. International Musculoskeletal Medicine 2015;37(1):29–32.

[39] McKibben LA, Iyer M, Zhao Y, Florea R, Kuhl-Chimera S, Deliwala I, Pan Y, Branham EM, Géranton SM, McLean SA, Linnstaedt SD. Transcriptional changes across tissue and time provide molecular insights into a therapeutic window of opportunity following traumatic stress exposure. bioRxiv 2024([Preprint]).

[40] McLean SA, Ressler K, Koenen KC, Neylan T, Germine L, Jovanovic T, Clifford GD, Zeng D, An X, Linnstaedt S, Beaudoin F, House S, Bollen KA, Musey P, Hendry P, Jones CW, Lewandowski C, Swor R, Datner E, Mohiuddin K, Stevens JS, Storrow A, Kurz MC, McGrath ME, Fermann GJ, Hudak LA, Gentile N, Chang AM, Peak DA, Pascual JL, Seamon MJ, Sergot P, Peacock WF, Diercks D, Sanchez LD, Rathlev N, Domeier R, Haran JP, Pearson C, Murty VP, Insel TR, Dagum P, Onnela J-P, Bruce SE, Gaynes BN, Joormann J, Miller MW, Pietrzak RH, Buysse DJ, Pizzagalli DA, Rauch SL, Harte SE, Young LJ, Barch DM, Lebois LAM, van Rooij SJH, Luna B, Smoller JW, Dougherty RF, Pace TWW, Binder E, Sheridan JF, Elliott JM, Basu A, Fromer M, Parlikar T, Zaslavsky AM, Kessler R. The AURORA Study: a longitudinal, multimodal library of brain biology and function after traumatic stress exposure. Molecular Psychiatry 2019.

[41] Michopoulos V, Powers A, Gillespie CF, Ressler KJ, Jovanovic T. Inflammation in Fear- and Anxiety-Based Disorders: PTSD, GAD, and Beyond. Neuropsychopharmacology 2017;42(1):254–270.

[42] Mills SEE, Nicolson KP, Smith BH. Chronic pain: a review of its epidemiology and associated factors in population-based studies. Br J Anaesth 2019;123(2):e273–e283.

[43] Mogil JS. Sex differences in pain and pain inhibition: multiple explanations of a controversial phenomenon. Nature Reviews Neuroscience 2012;13(12):859–866.

[44] Mogil JS. Qualitative sex differences in pain processing: emerging evidence of a biased literature. Nat Rev Neurosci 2020;21(7):353–365.

[45] Musey PI, Linnstaedt SD, Platts Mills TF, Miner JR, Bortsov AV, Safdar B, Bijur P, Rosenau A, Tsze DS, Chang AK. Gender differences in acute and chronic pain in the emergency department: results of the 2014 Academic Emergency Medicine consensus conference pain section. Academic Emergency Medicine 2014;21(12):1421–1430.

[46] Nehring SM, Goyal A, Patel BC. C Reactive Protein. StatPearls. Treasure Island (FL): StatPearls Publishing Copyright © 2024, StatPearls Publishing LLC., 2024.

[47] Nielsen CS. Assessing the societal cost of chronic pain. Scand J Pain 2022;22(4):684–685.

[48] Nievergelt CM, Maihofer AX, Klengel T, Atkinson EG, Chen CY, Choi KW, Coleman JRI, Dalvie S, Duncan LE, Gelernter J, Levey DF, Logue MW, Polimanti R, Provost AC, Ratanatharathorn A, Stein MB, Torres K, Aiello AE, Almli LM, Amstadter AB, Andersen SB, Andreassen OA, Arbisi PA, Ashley-Koch AE, Austin SB, Avdibegovic E, Babic D, Baekvad-Hansen M, Baker DG, Beckham JC, Bierut LJ, Bisson JI, Boks MP, Bolger EA, Borglum AD, Bradley B, Brashear M, Breen G, Bryant RA, Bustamante AC, Bybjerg-Grauholm J, Calabrese JR, Caldas-de-Almeida JM, Dale AM, Daly MJ, Daskalakis NP, Deckert J, Delahanty DL, Dennis MF, Disner SG, Domschke K, Dzubur-Kulenovic A, Erbes CR, Evans A, Farrer LA, Feeny NC, Flory JD, Forbes D, Franz CE, Galea S, Garrett ME, Gelaye B, Geuze E, Gillespie C, Uka AG, Gordon SD, Guffanti G, Hammamieh R, Harnal S, Hauser MA, Heath AC, Hemmings SMJ, Hougaard DM, Jakovljevic M, Jett M, Johnson EO, Jones I, Jovanovic T, Qin XJ, Junglen AG, Karstoft KI, Kaufman ML, Kessler RC, Khan A, Kimbrel NA, King AP, Koen N, Kranzler HR, Kremen WS, Lawford BR, Lebois LAM, Lewis CE, Linnstaedt SD, Lori A, Lugonja B, Luykx JJ, Lyons MJ, Maples-Keller J, Marmar C, Martin AR, Martin NG, Maurer D, Mavissakalian MR, McFarlane A, McGlinchey RE, McLaughlin KA, McLean SA, McLeay S, Mehta D, Milberg WP, Miller MW, Morey RA, Morris CP, Mors O, Mortensen PB, Neale BM, Nelson EC, Nordentoft M, Norman SB, O’Donnell M, Orcutt HK, Panizzon MS, Peters ES, Peterson AL, Peverill M, Pietrzak RH, Polusny MA, Rice JP, Ripke S, Risbrough VB, Roberts AL, Rothbaum AO, Rothbaum BO, Roy-Byrne P, Ruggiero K, Rung A, Rutten BPF, Saccone NL, Sanchez SE, Schijven D, Seedat S, Seligowski AV, Seng JS, Sheerin CM, Silove D, Smith AK, Smoller JW, Sponheim SR, Stein DJ, Stevens JS, Sumner JA, Teicher MH, Thompson WK, Trapido E, Uddin M, Ursano RJ, van den Heuvel LL, Van Hooff M, Vermetten E, Vinkers CH, Voisey J, Wang Y, Wang Z, Werge T, Williams MA, Williamson DE, Winternitz S, Wolf C, Wolf EJ, Wolff JD, Yehuda R, Young RM, Young KA, Zhao H, Zoellner LA, Liberzon I, Ressler KJ, Haas M, Koenen KC. International meta-analysis of PTSD genome-wide association studies identifies sex- and ancestry-specific genetic risk loci. Nat Commun 2019;10(1):4558.

[49] Osborne J. Improving your data transformations: Applying the Box-Cox transformation. Practical Assessment, Research, and Evaluation 2010;15(1):12.

[50] Otis JD, Gregor K, Hardway C, Morrison J, Scioli E, Sanderson K. An examination of the co-morbidity between chronic pain and posttraumatic stress disorder on US Veterans. Psychological Services 2010;7(3):126.

[51] Outcalt SD, Kroenke K, Krebs EE, Chumbler NR, Wu J, Yu Z, Bair MJ. Chronic pain and comorbid mental health conditions: independent associations of posttraumatic stress disorder and depression with pain, disability, and quality of life. Journal of behavioral medicine 2015;38(3):535–543.

[52] Paller CJ, Campbell CM, Edwards RR, Dobs AS. Sex-based differences in pain perception and treatment. Pain Med 2009;10(2):289–299.

[53] Parisien M, Lima LV, Dagostino C, El-Hachem N, Drury GL, Grant AV, Huising J, Verma V, Meloto CB, Silva JR, Dutra GGS, Markova T, Dang H, Tessier PA, Slade GD, Nackley AG, Ghasemlou N, Mogil JS, Allegri M, Diatchenko L. Acute inflammatory response via neutrophil activation protects against the development of chronic pain. Sci Transl Med 2022;14(644):eabj9954.

[54] Rivara FP, MacKenzie EJ, Jurkovich GJ, Nathens AB, Wang J, Scharfstein DO. Prevalence of Pain in Patients 1 Year After Major Trauma. Archives of Surgery 2008;143(3):282–287.

[55] Seligowski AV, Steuber ER, Hinrichs R, Reda MH, Wiltshire CN, Wanna CP, Winters SJ, Phillips KA, House SL, Beaudoin FL, An X, Stevens JS, Zeng D, Neylan TC, Clifford GD, Linnstaedt SD, Germine LT, Bollen KA, Guffanti G, Rauch SL, Haran JP, Storrow AB, Lewandowski C, Musey PI, Jr., Hendry PL, Sheikh S, Jones CW, Punches BE, Kurz MC, Murty VP, McGrath ME, Hudak LA, Pascual JL, Seamon MJ, Datner EM, Chang AM, Pearson C, Peak DA, Merchant RC, Domeier RM, Rathlev NK, O’Neil BJ, Sanchez LD, Bruce SE, Miller MW, Pietrzak RH, Joormann J, Barch DM, Pizzagalli DA, Sheridan JF, Luna B, Harte SE, Elliott JM, Koenen KC, Kessler RC, McLean SA, Ressler KJ, Jovanovic T. A prospective examination of sex differences in posttraumatic autonomic functioning. Neurobiol Stress 2021;15:100384.

[56] Shapiro SS, Wilk MB. An analysis of variance test for normality (complete samples). Biometrika 1965;52(3-4):591–611.

[57] Sorge RE, LaCroix-Fralish ML, Tuttle AH, Sotocinal SG, Austin J-S, Ritchie J, Chanda ML, Graham AC, Topham L, Beggs S. Spinal cord Toll-like receptor 4 mediates inflammatory and neuropathic hypersensitivity in male but not female mice. Journal of Neuroscience 2011;31(43):15450–15454.

[58] Sorge RE, Totsch SK. Sex differences in pain. Journal of neuroscience research 2017;95(6):1271–1281.

[59] Toblin RL, Quartana PJ, Riviere LA, Walper KC, Hoge CW. Chronic pain and opioid use in US soldiers after combat deployment. JAMA internal medicine 2014;174(8):1400–1401.

[60] Vongpatanasin W, Tuncel M, Wang Z, Arbique D, Mehrad B, Jialal I. Differential effects of oral versus transdermal estrogen replacement therapy on C-reactive protein in postmenopausal women. Journal of the American College of Cardiology 2003;41(8):1358–1363.

[61] Xu Z-Z, Zhang L, Liu T, Park JY, Berta T, Yang R, Serhan CN, Ji R-R. Resolvins RvE1 and RvD1 attenuate inflammatory pain via central and peripheral actions. Nature medicine 2010;16(5):592–597.

