## Supplemental Material for "Peritraumatic C-reactive protein levels predict pain outcomes following traumatic stress exposure in a sex-dependent manner"

***Supplementary Table 1***. Full model statistics for the relationship between peritraumatic (i.e. ED) CRP levels and longitudinal pain outcomes (week 8, month 3, month 6) in men and women

| | **Men (n=99)** | | | | **Women (n=220)** | | | |
|---|---|---|---|---|---|---|---|---|
| | **β** | **SEM** | **t-value** | ***p*** | **β** | **SEM** | **t-value** | ***p*** |
| **Time** | -0.36 | 0.08 | -4.71 | **<0.001** | -0.16 | 0.05 | -3.39 | **<0.001** |
| **ED CRP** | -0.24 | 0.11 | -2.13 | **0.037** | 0.05 | 0.07 | 0.72 | 0.470 |
| **Age** | 0.03 | 0.02 | 1.50 | 0.137 | 0.02 | 0.01 | 1.44 | 0.150 |
| **ED pain** | 0.38 | 0.10 | 3.70 | **<0.001** | 0.38 | 0.08 | 4.88 | **<0.001** |
| **Highest grade: high school** | -0.56 | 0.91 | -0.62 | 0.539 | -0.19 | 0.73 | -0.26 | 0.799 |
| **Highest grade: some college** | 0.15 | 0.83 | 0.18 | 0.855 | 0.21 | 0.66 | 0.31 | 0.756 |
| **Highest grade: ≥ bachelor's degree** | 0.17 | 1.04 | 0.16 | 0.872 | -0.09 | 0.74 | -0.12 | 0.904 |
| **Race** | 0.39 | 0.59 | 0.65 | 0.516 | 0.11 | 0.44 | 0.24 | 0.812 |
| **Plate 2** | -0.55 | 1.10 | -0.50 | 0.620 | 1.46 | 0.79 | 1.85 | 0.066 |
| **Plate 3** | 0.79 | 1.18 | 0.67 | 0.502 | 1.68 | 0.79 | 2.13 | **0.034** |
| **Plate 4** | 0.19 | 1.50 | 0.13 | 0.900 | 1.58 | 0.85 | 1.86 | 0.064 |
| **Plate 5** | -0.57 | 1.71 | -0.33 | 0.739 | 0.89 | 0.94 | 0.95 | 0.344 |
| **Plate 6** | -0.09 | 1.83 | -0.05 | 0.959 | 1.10 | 0.98 | 1.13 | 0.262 |
| **Plate 7** | -0.81 | 1.40 | -0.58 | 0.565 | 1.62 | 0.91 | 1.78 | 0.076 |
| **Plate 8** | 1.52 | 1.61 | 0.94 | 0.349 | 2.37 | 0.92 | 2.58 | **0.011** |
| **Plate 9** | 0.44 | 1.16 | 0.38 | 0.703 | 2.09 | 0.74 | 2.82 | **0.005** |
| **Plate 10** | -2.49 | 1.46 | -1.71 | 0.092 | 1.73 | 0.85 | 2.05 | 0.042 |
| **Plate 11** | 3.15 | 2.06 | 1.53 | 0.130 | 0.81 | 1.21 | 0.67 | 0.503 |
| **Plate 12** | 0.72 | 1.38 | 0.52 | 0.607 | 1.31 | 0.86 | 1.53 | 0.129 |
| **Plate 13** | -2.28 | 1.79 | -1.27 | 0.208 | 2.24 | 1.12 | 2.00 | 0.047 |

ED: emergency department, CRP: C-reactive protein, SEM: standard error of the mean
CRP was $log_2$-adjusted

***Supplementary Table 2.*** Full model statistics for the relationship between peritraumatic (i.e. ED) CRP levels and longitudinal pain outcomes (week 8, month 3, month 6) adjusted for BMI, in men and women

| | **Men (n=78)** | | | | **Women (n=197)** | | | |
|---|---|---|---|---|---|---|---|---|
| | **β** | **SEM** | **t-value** | ***p*** | **β** | **SEM** | **t-value** | ***p*** |
| **Time** | -0.35 | 0.08 | -4.32 | **<0.001** | -0.14 | 0.05 | -2.77 | **0.006** |
| **CRP** | -0.23 | 0.14 | -1.63 | **0.110** | 0.04 | 0.09 | 0.46 | 0.643 |
| **Age** | 0.01 | 0.02 | 0.36 | 0.720 | 0.02 | 0.02 | 1.29 | 0.199 |
| **ED pain** | 0.24 | 0.12 | 1.93 | **0.059** | 0.36 | 0.08 | 4.38 | **<0.001** |
| **Highest grade: high school** | -0.54 | 1.12 | -0.48 | 0.630 | 0.14 | 0.76 | 0.19 | 0.850 |
| **Highest grade: some college** | 0.13 | 1.00 | 0.13 | 0.899 | 0.17 | 0.69 | 0.24 | 0.808 |
| **Highest grade: ≥ bachelor's degree** | -0.42 | 1.27 | -0.33 | 0.743 | 0.06 | 0.76 | 0.08 | 0.933 |
| **Race** | 0.29 | 0.70 | 0.42 | 0.679 | 0.29 | 0.47 | 0.60 | 0.547 |
| **BMI** | 0.00 | 0.05 | -0.02 | 0.987 | 0.03 | 0.03 | 0.86 | 0.392 |
| **Plate 2** | 0.74 | 1.50 | 0.49 | 0.623 | 1.61 | 0.87 | 1.85 | 0.066 |
| **Plate 3** | 0.54 | 1.75 | 0.31 | 0.759 | 2.19 | 0.87 | 2.52 | **0.013** |
| **Plate 4** | 2.87 | 1.98 | 1.45 | 0.152 | 1.76 | 0.94 | 1.86 | 0.064 |
| **Plate 5** | 0.07 | 1.95 | 0.04 | 0.970 | 0.90 | 0.99 | 0.91 | 0.365 |
| **Plate 6** | 0.91 | 2.45 | 0.37 | 0.713 | 1.30 | 1.09 | 1.19 | 0.236 |
| **Plate 7** | -0.61 | 1.87 | -0.33 | 0.745 | 1.78 | 1.01 | 1.76 | 0.080 |
| **Plate 8** | 2.45 | 1.89 | 1.30 | 0.201 | 2.44 | 0.96 | 2.54 | **0.012** |
| **Plate 9** | 1.80 | 1.54 | 1.17 | 0.248 | 1.93 | 0.78 | 2.48 | **0.014** |
| **Plate 10** | -1.21 | 1.78 | -0.68 | 0.499 | 1.62 | 0.88 | 1.83 | 0.068 |
| **Plate 11** | 4.27 | 2.31 | 1.85 | 0.070 | 0.68 | 1.23 | 0.55 | 0.584 |
| **Plate 12** | 2.05 | 1.71 | 1.20 | 0.235 | 1.28 | 0.90 | 1.43 | 0.156 |
| **Plate 13** | -1.27 | 2.02 | -0.63 | 0.533 | 2.15 | 1.15 | 1.86 | 0.065 |

ED:emergency department, CRP: C-reactive protein, BMI: body mass index, SEM: standard error of the mean
CRP was $log_2$-adjusted

***Supplementary Table 3.*** Full model statistics for the relationship between peritraumatic (i.e. ED) CRP levels and longitudinal pain outcomes (week 8, month 3, month 6) in men and women, the subcohort of participants with available BMI data (BMI not included as a covariate)

| | **Men (n=78)** | | | | **Women (n=197)** | | | |
|---|---|---|---|---|---|---|---|---|
| | **β** | **SEM** | **t-value** | ***p*** | **β** | **SEM** | **t-value** | ***p*** |
| **Time** | -0.35 | 0.08 | -4.32 | **<0.001** | -0.14 | 0.05 | -2.77 | **0.006** |
| **CRP** | -0.23 | 0.14 | -1.69 | **0.096** | 0.07 | 0.08 | 0.94 | 0.346 |
| **Age** | 0.01 | 0.02 | 0.37 | 0.716 | 0.02 | 0.01 | 1.42 | 0.159 |
| **ED pain** | 0.24 | 0.12 | 1.95 | **0.056** | 0.37 | 0.08 | 4.58 | **<0.001** |
| **Highest grade: high school** | -0.54 | 1.11 | -0.49 | 0.627 | 0.15 | 0.76 | 0.20 | 0.842 |
| **Highest grade: some college** | 0.13 | 0.99 | 0.13 | 0.898 | 0.23 | 0.69 | 0.33 | 0.743 |
| **Highest grade: ≥ bachelor's degree** | -0.42 | 1.23 | -0.34 | 0.732 | 0.06 | 0.76 | 0.08 | 0.935 |
| **Race** | 0.29 | 0.68 | 0.42 | 0.674 | 0.30 | 0.47 | 0.63 | 0.526 |
| **Plate 2** | 0.75 | 1.46 | 0.51 | 0.611 | 1.52 | 0.86 | 1.76 | 0.080 |
| **Plate 3** | 0.55 | 1.66 | 0.33 | 0.742 | 2.17 | 0.87 | 2.50 | **0.014** |
| **Plate 4** | 2.87 | 1.96 | 1.47 | 0.148 | 1.70 | 0.94 | 1.81 | 0.072 |
| **Plate 5** | 0.08 | 1.88 | 0.04 | 0.966 | 0.83 | 0.99 | 0.83 | 0.405 |
| **Plate 6** | 0.91 | 2.41 | 0.38 | 0.706 | 1.23 | 1.09 | 1.14 | 0.258 |
| **Plate 7** | -0.61 | 1.86 | -0.33 | 0.742 | 1.71 | 1.01 | 1.70 | 0.091 |
| **Plate 8** | 2.46 | 1.87 | 1.32 | 0.194 | 2.31 | 0.95 | 2.44 | **0.016** |
| **Plate 9** | 1.81 | 1.50 | 1.20 | 0.234 | 1.95 | 0.78 | 2.50 | **0.013** |
| **Plate 10** | -1.21 | 1.74 | -0.70 | 0.490 | 1.64 | 0.88 | 1.86 | 0.065 |
| **Plate 11** | 4.27 | 2.29 | 1.87 | 0.067 | 0.68 | 1.23 | 0.55 | 0.581 |
| **Plate 12** | 2.06 | 1.67 | 1.23 | 0.224 | 1.25 | 0.90 | 1.40 | 0.165 |
| **Plate 13** | -1.27 | 2.00 | -0.63 | 0.529 | 2.18 | 1.15 | 1.89 | 0.061 |

ED:emergency department, CRP: C-reactive protein, BMI: body mass index, SEM: standard error of the mean
CRP was $log_2$-adjusted

***Supplementary Table 4*. Counts of participants with increasing versus decreasing CRP and Pain trends.**

| | | **CRP Trend from ED to 6M** | | | |
|---|---|---|---|---|---|
| | | **Decreasing CRP** | | **Increasing CRP** | |
| | | **Men** | **Women** | **Men** | **Women** |
| **Pain Trend from ED to 6M** | **Decreasing Pain** | n= 39 | n=72 | n=27 | n=62 |
| | **Increasing Pain** | n=6 | n=28 | n=1 | n=26 |
| | **Unchanged** | n=2 | n=10 | n=4 | n=14 |

Men: χ2-test *p*=0.115
Women: χ2-test *p*=0.552
CRP: C-reactive protein, ED: emergency department, 6M: six months
